# Association of renal function with diabetic retinopathy and macular edema among patients with type 2 diabetes mellitus

**DOI:** 10.1101/2020.11.22.20236265

**Authors:** Lanhua Wang, Ling Jin, Wei Wang, Xia Gong, Yuting Li, Wangting Li, Jie Meng, Xiaoling Liang, Wenyong Huang, Yizhi Liu

## Abstract

**Purpose:** To investigate the associations between renal function and the presence of diabetic retinopathy (DR) in diabetic patients.

**Methods:** A total of 1877 diabetic participants aged 30 to 80 years were consecutively recruited from October 2017 to April 2019. All participants underwent blood and urine analyses and standardized 7-field fundus imaging. The presence of DR, vision-threatening DR (VTDR) and DME was graded based on the fundus photographs. Renal function was defined as normal, mildly impaired or chronic kidney disease (CKD) based on different estimated glomerular filtration rates (GFRs).

**Results:** Unlike a normal GFR, CKD was significantly associated with any DR (OR=1.89, P=0.017) and VTDR (OR=2.76, P=0.009), and mildly impaired renal function was associated with only any DR (OR=1.39, P=0.031). The analysis of the effect of microalbuminuria on relationship between GFR and DR showed that the GFR was associated with any DR only in the presence of microalbuminuria, while the GFR was an independent risk factor for VTDR regardless of microalbuminuria status (all P<0.05). The risks of any DR (OR=1.74 for quartile 2 and 3.09 for quartile 4) and VTDR (OR=3.27 for quartile 2 and 6.41 for quartile 4) increased gradually as the microalbuminuria quartile increased (all P<0.05). The third (OR=2.99, P=0.029) and fourth microalbuminuria quartiles (OR=4.74, P=0.002) were independent DME risk factors.

**Conclusions:** There was a strong association between GFR and VTDR, whereas the association of GFR and any DR was significant only under the premise of microalbuminuria. High microalbuminuria was significantly associated with DR and DME.

## Introduction

The number of people with diabetes mellitus (DM) is increasing worldwide, resulting in an increase in prevalent diabetic microvascular complications, which mainly include diabetic retinopathy (DR) and diabetic nephropathy (DN).^1^ DR is a leading cause of vision impairment and blindness in working-aged people.^2-5^ It is estimated that without prompt treatment, the number of people with DR will increase rapidly from 126.6 million in 2010 to 191 million by 2030 worldwide.^6^ Progressive microvascular damage is also commonly observed in glomeruli which may lead to microalbuminuria, a decrease in the glomerular filtration rate (GFR) and the eventual development of chronic kidney disease (CKD).^7^

There is an increasing interesting in the literature about the relationship between DR and DN due to the shared risk factors and similar pathophysiological features.^8, 9^ Numerous studies have consistently demonstrated a strong association between DN and DR in type 1 diabetes mellitus (T1DM) patients, but this association is not as strong in type 2 diabetes mellitus (T2DM) patients.^10, 11^ Recent studies on the relationship between the GFR and DR among T2DM patients showed equivocal results, with some studies indicating a negative association between the GFR and DR,^10, 11^ others showing no association.^12^

The role of microalbuminuria as a biomarker for microvascular damage of DM is debatable because some studies have indicated that microalbuminuria may regress to normal over time.^13, 14^ Additionally, studies found a dissociated relationship between GFRs and microalbuminuria levels in DM patients^15^ and about 50% of T2DM patients ^16, 17^ with normal microalbuminuria levels had decreased GFR levels, possibly due to nondiabetic causes^18^, aging^13^, kidney damage, or aggressive antihypertensive treatment.^16, 19^ Studies investigating whether the GFR and microalbuminuria are competing risk factors for DR are limited and controversial.^12, 20^

Therefore, we conducted a cross-sectional study to investigate the association of the GFR with the presence of DR and diabetic macular edema (DME) in Chinese patients with T2DM. The effect of microalbuminuria on the association of the GFR with DR was also assessed.

## Methods

### Study population

This cross-sectional study was conducted from October 2017 to April 2019 at the Zhongshan Ophthalmic Center (ZOC), Guangzhou, Southern China.

Individuals with T2DM aged between 30 and 80 years who were registered in the community health system near ZOC were consecutively recruited to participate in the study. Subjects were not recruited if any of the following conditions were present: (1) a previous history of cerebrovascular disease, cardiovascular disease, malignancy or other severe systemic disease that were not conducive to comprehensive examination; (2) pregnancy; (3) cognitive disorders or mental diseases; (4) ungradable bilateral fundus photographs; or (5) refusal of or intolerability to mydriasis due to shallow anterior chamber or angle closure glaucoma. The study adhered to the tenets of the Declaration of Helsinki, and approval was obtained from the Institutional Ethical Committee of ZOC. Written informed consent was obtained from all participants.

### Study procedure

#### Assessment of diabetic retinopathy and diabetic macular edema

Except for those with a van Herick value of less than 25° or an intraocular pressure (IOP) of 21 mmHg or more, all participants had their pupils dilated with 0.5% topicamide plus 0.5% phenylephrine eye drop. Then, standardized 7-field fundus photographs using a digital camera (Canon CR-2, Tokyo, Japan) were taken 30 minutes after mydriasis. Fundus photographs were independently graded by 2 trained graders according to the United Kingdom National Diabetic Eye Screening Program (UK NDESP) guidelines.^21^

According to the UK NDESP guidelines, R0 was defined as no diabetic retinal lesion. R1 was defined as the presence of at least one of the following retinal lesions: microaneurysm, dot hemorrhages, hard exudate (HE), cotton wool spots, and venous loops. R2 was defined as the presence of any retinal lesion including venous beading, venous reduplication, multiple blot hemorrhage, or intraretinal microvascular abnormalities. R3 was considered present with the presence of new vessels on the disc retina or elsewhere; preretinal fibrosis, preretinal or vitreous hemorrhage, or evidence of previous peripheral retinal laser treatment.^21^

DME was defined as the presence of HE within 1 disc diameter of the fovea center, a collection of exudates within the macula, or the presence of focal photocoagulation scars in the macular area. Participants with any presence of R2, R3 or DME were considered to have vision-threatening diabetic retinopathy (VTDR).

#### Assessment of blood and urine chemistry

A venous blood sample was collected to estimate serum uric acid (UA), the serum creatinine clearance rate (CCr), C-reactive protein (CPR), total cholesterol (TC), triglycerides (TGs), low-density lipoprotein cholesterol (LDL-c), and high-density lipoprotein cholesterol (HDL-c) using a chemical analyzer (Cobs 8000; Roche Diagnostics; Mannheim, Germany). Hemoglobin A1C (HbA1c) was tested by a Sysmex G8 glycosylated hemoglobin analyzer (Sysmex Corporation; Kobe, Japan) at the clinical laboratory of ZOC. A 50-ml midstream urine sample was collected to assess microalbuminuria with a Cobs 8000 biochemistry analyzer (Roche Diagnostics; Mannheim, Germany). The estimated GFR was calculated according to the widely used Chronic Kidney Disease Epidemiology Collaboration (CKD-EPI) equation.^22^ Renal function was defined as normal (GFR > 90 mL/min/1.73 m^2^), mildly impaired (GFR 60-89 mL/min/1.73 m^2^), or CKD (GFR < 60 mL/min/1.73 m^2^).^23^

#### Assessment of other examinations

All participants underwent other ocular examinations, including visual acuity, IOP, auto-refraction, ocular biometry, and slit lamp examination of anterior and posterior segment. Systolic (SBP) and diastolic (DBP) blood pressures, height and weight were measured by a trained nurse with a standard operating protocol. Mean arterial pressure (MAP) was defined as one-third of SBP plus two-thirds of DBP. Body mass index (BMI) was calculated as the weight(kg) divided by the square of the height(m). A standard questionnaire was administered by a trained interviewer to collect information on demography, lifestyle risk factors, systemic and eye medical histories.

### Statistical analysis

DR was graded based on the worse eye in those with bilateral qualified fundus images. All statistical analyses were performed with Stata (ver. 12.0; Stata Corp; College Station, TX). Student’s t test or one-way ANOVA was used to compare continuous variables, while the chi-squared test was used to compare categorical data. Multiple logistic regression models were used to test the association between the presence of DR, VTDR, or DME and renal function (microalbuminuria and GFR) after adjusting for confounders. Odds ratios (OR) and 95% confidence intervals (CI) are presented. A P value of < 0.05 was considered to be statistically significant.

## Results

A total of 1877 participants with T2DM were enrolled in the current study. Of the 1877 eligible participants, 380 (20.3%) had any DR, and 66 (3.52%) had DME. Participants with any DR were less likely to be male (P=0.005), had a longer DM duration (P<0.001), were more likely to use insulin (P<0.001), were more likely to have a history of hyperlipidemia (P<0.001), had higher HbA1c (P<0.001), had higher SBP (P<0.001), had higher GFR (P<0.001), and had a higher microalbuminuria level (P<0.001) than those without any DR. Participants with DME tended to be younger (P=0.001), more likely to use insulin (P=0.001), less likely to have a history of hyperlipidemia (P<0.001), had higher HbA1c (P<0.001), had higher SBP (P<0.001), and had higher LDL-C (P=0.047) than those without DME (Table 1).

**Table1.** Comparison of basic characteristic of participants with and without diabetic retinopathy

| Characteristic | DR |  |  | DME |  |  |
| --- | --- | --- | --- | --- | --- | --- |
|  | No DR | Any DR | P-value | No DME | DME | P-value |
| Total number, n (%) | 1497(79.8) | 380(20.3) |  | 1811(96.5) | 66(3.52) |  |
| Age(years) | 64.6±7.63 | 63.8±8.14 | 0.060 | 64.6±7.63 | 61.1±9.78 | 0.001 |
| Female, n (%) | 888(59.3) | 195(51.3) | 0.005 | 1049(57.9) | 34(51.5) | 0.301 |
| Duration of diabetes(years) | 8.24±6.67 | 11.8±7.72 | <0.001 | 8.91±7.05 | 10.6±6.65 | 0.056 |
| Use of insulin, n (%) | 325(21.8) | 167(44.1) | <0.001 | 463(25.7) | 29(43.9) | 0.001 |
| Current smoker, n (%) | 215(25.3) | 53(26.1) | 0.811 | 257(25.4) | 11(27.5) | 0.762 |
| Current drinker, n (%) | 207(24.4) | 53(26.1) | 0.602 | 249(24.6) | 11(27.5) | 0.674 |
| History of hypertension,n (%) | 855(57.1) | 218(57.4) | 0.929 | 1033(57.0) | 40(60.6) | 0.565 |
| History of hyperlipidemia,n (%) | 730(48.8) | 145(38.2) | <0.001 | 857(47.3) | 18(27.3) | <0.001 |
| HbA1c(%) | 6.77±1.27 | 7.73±1.75 | <0.001 | 6.92±1.40 | 8.14±1.65 | <0.001 |
| BMI (kg/m<sup>2</sup>) | 24.7±3.26 | 24.4±3.30 | 0.062 | 24.7±3.27 | 24.1±3.36 | 0.171 |
| MAP (mmHg) | 91.5±11.8 | 93.2±12.2 | 0.011 | 91.8±11.9 | 93.3±13.1 | 0.313 |
| GFR (ml/min/1.73m<sup>2</sup>) | 85.6±16.4 | 81.3±19.6 | <0.001 | 84.8±17.0 | 82.8±21.5 | 0.348 |
| Cholesterol (mg/dL) | 185.9±39.9 | 186.2±42.1 | 0.888 | 185.7±40.1 | 193.6±47.2 | 0.116 |
| LDL-C(mg/dL) | 118.2±36.3 | 120.0±36.6 | 0.371 | 118.2±36.1 | 127.3±42.8 | 0.047 |
| HDL-C(mg/dL) | 50.6±15.6 | 49.9±15.6 | 0.416 | 50.5±15.6 | 49.7±15.3 | 0.701 |
| Triglycerides (mg/dL) | 206.4±143.0 | 206.4±153.1 | 0.999 | 206.2±145.3 | 210.8±138.0 | 0.802 |
| UA (mg/dL) | 6.20±1.62 | 6.12±1.74 | 0.417 | 6.19±1.64 | 6.06±1.72 | 0.554 |
| CPR(mg/dL) | 27.6±69.8 | 26.6±49.3 | 0.789 | 27.5±67.2 | 23.1±21.5 | 0.591 |
| Microalbuminuria(mg/dL) | 4.03±11.9 | 15.3±63.1 | <0.001 | 5.99±30.4 | 13.3±23.2 | 0.065 |

All data presented as mean±standard deviation unless otherwise indicated
Abbreviations: DR = diabetic retinopathy; DME = diabetic macular edema;
HbA1c = hemoglobin A1c; BMI = body mass index; MAP =mean arterial
pressure; GFR = glomerular filtration rate; LDL-C=low density lipoprotein;
HDL-C=high density lipoprotein; UA=uric acid; CPR=C-reactive protein

In general, participants with impaired renal function were more likely to be older (P<0.001), were more likely to be male (P<0.001), had a longer DM duration (P<0.001), were more likely to have a history of hypertension (P<0.001), had a higher BMI (P=0.016), had higher UA (P<0.001), had higher CPR (P=0.009), had a higher microalbuminuria level (P<0.001), and had higher risks of any DR (P=0.012) and VTDR (P=0.006) than those with normal GFR. (Table 2).

**Table2.**
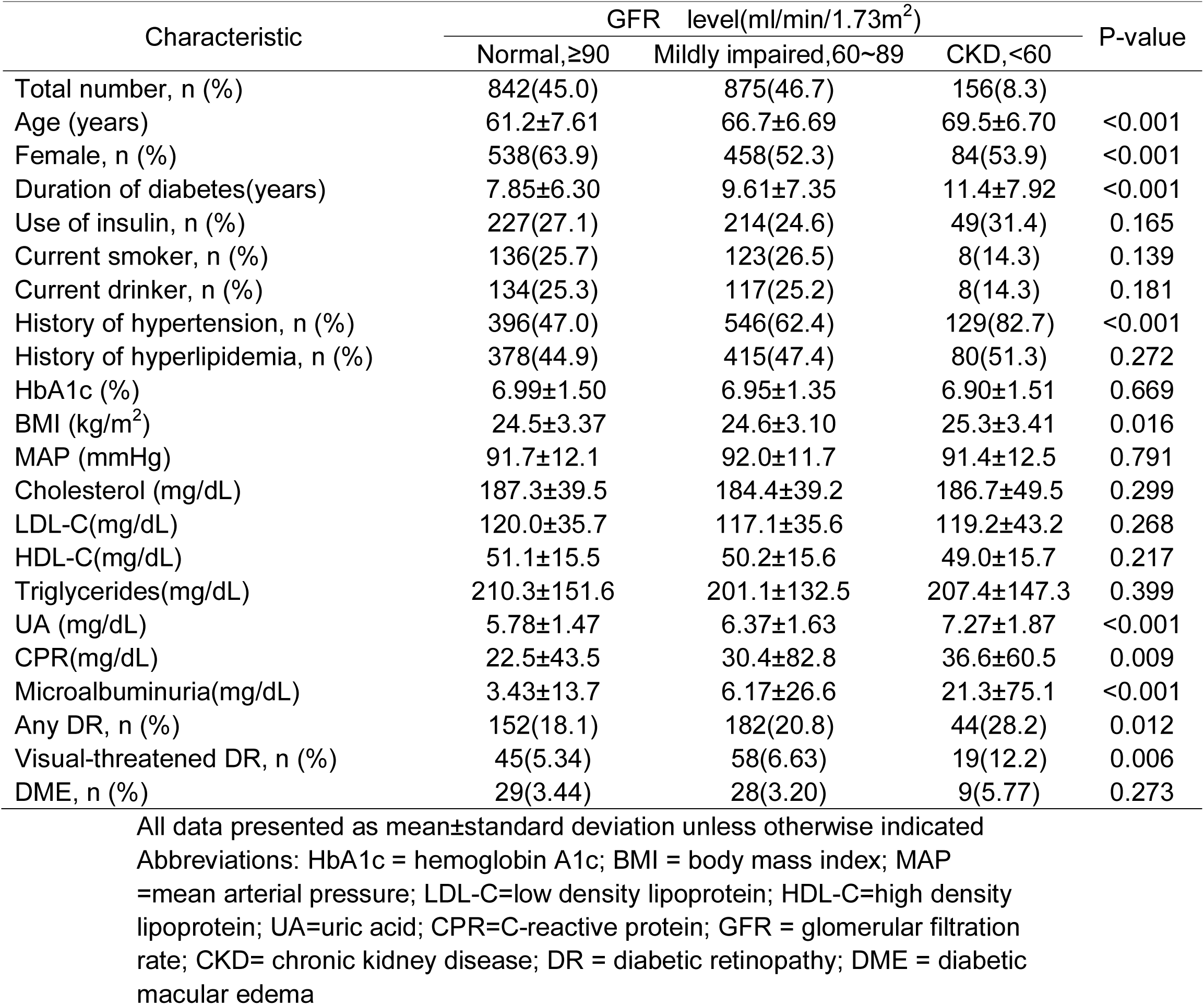
Distribution of basic characteristic stratified by GFR among participants with diabetes

| Characteristic | GFR level(ml/min/1.73m<sup>2</sup>) |  |  | P-value |
| --- | --- | --- | --- | --- |
|  | Normal, ≥90 | Mildly impaired, 60~89 | CKD, <60 |  |
| Total number, n (%) | 842(45.0) | 875(46.7) | 156(8.3) |  |
| Age (years) | 61.2±7.61 | 66.7±6.69 | 69.5±6.70 | <0.001 |
| Female, n (%) | 538(63.9) | 458(52.3) | 84(53.9) | <0.001 |
| Duration of diabetes(years) | 7.85±6.30 | 9.61±7.35 | 11.4±7.92 | <0.001 |
| Use of insulin, n (%) | 227(27.1) | 214(24.6) | 49(31.4) | 0.165 |
| Current smoker, n (%) | 136(25.7) | 123(26.5) | 8(14.3) | 0.139 |
| Current drinker, n (%) | 134(25.3) | 117(25.2) | 8(14.3) | 0.181 |
| History of hypertension, n (%) | 396(47.0) | 546(62.4) | 129(82.7) | <0.001 |
| History of hyperlipidemia, n (%) | 378(44.9) | 415(47.4) | 80(51.3) | 0.272 |
| HbA1c (%) | 6.99±1.50 | 6.95±1.35 | 6.90±1.51 | 0.669 |
| BMI (kg/m<sup>2</sup>) | 24.5±3.37 | 24.6±3.10 | 25.3±3.41 | 0.016 |
| MAP (mmHg) | 91.7±12.1 | 92.0±11.7 | 91.4±12.5 | 0.791 |
| Cholesterol (mg/dL) | 187.3±39.5 | 184.4±39.2 | 186.7±49.5 | 0.299 |
| LDL-C(mg/dL) | 120.0±35.7 | 117.1±35.6 | 119.2±43.2 | 0.268 |
| HDL-C(mg/dL) | 51.1±15.5 | 50.2±15.6 | 49.0±15.7 | 0.217 |
| Triglycerides(mg/dL) | 210.3±151.6 | 201.1±132.5 | 207.4±147.3 | 0.399 |
| UA (mg/dL) | 5.78±1.47 | 6.37±1.63 | 7.27±1.87 | <0.001 |
| CPR(mg/dL) | 22.5±43.5 | 30.4±82.8 | 36.6±60.5 | 0.009 |
| Microalbuminuria(mg/dL) | 3.43±13.7 | 6.17±26.6 | 21.3±75.1 | <0.001 |
| Any DR, n (%) | 152(18.1) | 182(20.8) | 44(28.2) | 0.012 |
| Visual-threatened DR, n (%) | 45(5.34) | 58(6.63) | 19(12.2) | 0.006 |
| DME, n (%) | 29(3.44) | 28(3.20) | 9(5.77) | 0.273 |

All data presented as mean±standard deviation unless otherwise indicated
Abbreviations: HbA1c = hemoglobin A1c; BMI = body mass index; MAP =mean arterial pressure; LDL-C=low density lipoprotein; HDL-C=high density lipoprotein; UA=uric acid; CPR=C-reactive protein; GFR = glomerular filtration rate; CKD= chronic kidney disease; DR = diabetic retinopathy; DME = diabetic macular edema

Participants in the higher quartile of microalbuminuria level tended to be older (P<0.001), were less likely to be female (P<0.001), had a longer DM duration (P<0.001), had higher HbA1c (P<0.001), were more likely to use insulin (P<0.001), were more likely to have history of hypertension (P<0.001), had a higher BMI (P<0.001), had a higher MAP (P=0.015), had a lower GFR (P<0.001), had lower HDL-c (P<0.001), had higher TGs (P<0.001), had higher UA (P<0.001), had higher CRP (P=0.002), had higher risks of any DR (P<0.001), VTDR (P<0.001) and DME (P<0.001) than those in the first quartile(Table 3).

**Table3.** Distribution of basic characteristic stratified by microalbuminuria level among participants with diabetes

| Characteristic | Microalbuminuria level(mg/dL) |  |  |  | P value |
| --- | --- | --- | --- | --- | --- |
|  | First quartile (<22.85) | Second quartile (22.85~23.43) | Third quartile (23.43~24.06) | Fourth quartile (≥24.06) |  |
| Age (years) | 63.3±7.60 | 65.1±7.44 | 65.1±8.24 | 66.4±8.25 | <0.001 |
| Female, n (%) | 308(65.1) | 273(61.6) | 256(54.9) | 239(50.3) | <0.001 |
| Duration of diabetes(years) | 7.85±6.26 | 8.95±6.98 | 8.64±6.61 | 10.2±7.80 | <0.001 |
| HbA1c (%) | 6.80±1.34 | 6.86±1.40 | 6.90±1.40 | 7.25±1.57 | <0.001 |
| Use of insulin, n (%) | 101(21.49) | 107(24.4) | 103(22.2) | 160(33.9) | <0.001 |
| Current smoker, n (%) | 80(20.2) | 55(23.1) | 55(26.3) | 49(25.0) | 0.322 |
| Current drinker, n (%) | 75(18.9) | 50(21.0) | 46(22.0) | 51(26.0) | 0.263 |
| History of hypertension, n (%) | 216(46.6) | 239(54.1) | 283(61.4) | 344(73.4) | <0.001 |
| History of hyperlipidemia, n (%) | 218(46.7) | 198(45.4) | 210(46.6) | 241(52.5) | 0.134 |
| BMI (kg/m<sup>2</sup>) | 24.0±2.99 | 24.2±3.19 | 24.9±3.34 | 25.4±3.55 | <0.001 |
| MAP(mmHg) | 90.4±11.1 | 90.1±10.8 | 91.8±12.0 | 94.8±12.9 | <0.001 |
| GFR (ml/min/1.73m<sup>2</sup>) | 90.5±13.7 | 86.6±16.2 | 84.6±16.3 | 76.5±19.4 | <0.001 |
| Cholesterol(mg/dL) | 186.1±39.8 | 183.5±39.9 | 183.2±39.6 | 186.8±43.4 | 0.427 |
| LDL-C(mg/dL) | 118.2±35.4 | 117.5±35.6 | 116.536.3± | 118.6±38.8 | 0.837 |
| HDL-C(mg/dL) | 52.6±16.7 | 51.8±15.4 | 49.9±15.7 | 47.2±14.7 | <0.001 |
| Triglycerides(mg/dL) | 197.1±139.1 | 186.9±121.4 | 207.3±137.7 | 233.8±164.6 | <0.001 |
| UA(mg/dL) | 5.98±1.52 | 6.02±1.63 | 6.24±1.50 | 6.56±1.83 | <0.001 |
| CPR(mg/dL) | 23.0±55.9 | 23.5±42.9 | 27.3±45.1 | 38.2±105.3 | 0.002 |
| Any DR, n (%) | 48(10.5) | 75(17.4) | 84(18.9) | 142(31.9) | <0.001 |
| Visual-threatened DR, n (%) | 8(1.74) | 23(5.35) | 26(5.84) | 56(12.6) | <0.001 |
| DME, n (%) | 6(1.31) | 9(2.09) | 17(3.82) | 28(6.29) | <0.001 |

All data presented as mean±standard deviation unless otherwise indicated
Abbreviations: HbA1c = hemoglobin A1c; BMI = body mass index; MAP =mean arterial pressure; GFR = glomerular filtration rate; LDL-C=low density lipoprotein; HDL-C=high density lipoprotein; UA=uric acid; CPR=C-reactive protein; DR = diabetic retinopathy; DME = diabetic macular edema

In multiple logistic regression model, a low GFR was significantly associated with the presence of DR(OR=0.98 per 1 mL/min/1.73 m^2^ increase, P<0.001) and VTDR (OR=0.98 per 1 mL/min/1.73 m^2^ increase, P<0.001).We also assessed the associations of DR with different GFR levels, and the results showed that CKD were an important risk factor for any DR (OR=1.89, P=0.017) and VTDR (OR=2.76, P=0.009) compared to a normal GFR, and mildly impaired renal function (OR=1.39, P=0.031) was significantly associated with any DR. A high microalbuminuria level was significantly associated with any DR (OR=1.01 per 1 mg/dl increase, P<0.001) and VTDR (OR=1.39 per 1 mg/dl increase, P=0.003) in the multiple logistic regression model, and participants in the fourth quartile had a 3.09-fold (P<0.001) higher risk for any DR and a 6.41-fold (P<0.001) higher risk for VTDR than participants in the first microalbuminuria quartile (Table 4).

**Table 4.** Logistic regression of GFR and microalbuminuria with presence of any DR and VTDR

| Factors | Any DR |  |  |  | VTDR |  |  |  |
| --- | --- | --- | --- | --- | --- | --- | --- | --- |
|  | Model 1<sup>*</sup> |  | Model 2<sup>†</sup> |  | Model 1<sup>*</sup> |  | Model 2<sup>†</sup> |  |
|  | OR(95%CI) | P value | OR(95%CI) | P value | OR(95%CI) | P value | OR(95%CI) | P value |
| **GFR(ml/min/1.73m<sup>2</sup>)** |  |  |  |  |  |  |  |  |
| Normal, ≥90 | ref | ref | ref | ref | ref | ref | ref | ref |
| Mildly impaired, 60~89 | 1.31(1.01,1.70) | 0.039 | 1.39(1.03,1.88) | 0.031 | 1.70(1.10 , 2.63) | 0.018 | 1.40(0.85,2.32) | 0.191 |
| CKD, <60 | 2.11(1.39,3.20) | <0.001 | 1.89(1.12,3.17) | 0.017 | 3.97(2.14 , 7.39) | <0.001 | 2.76(1.29,5.92) | 0.009 |
| Per 1-ml/min/1.73m<sup>2</sup> increased | 0.98(0.97,0.99) | <0.001 | 0.98(0.97,0.99) | <0.001 | 0.97(0.95,0.98) | <0.001 | 0.98(0.97,0.99) | 0.001 |
| **Microalbuminuria(mg/dL)** |  |  |  |  |  |  |  |  |
| Quartiles1 | ref | ref | ref | ref | ref | ref | ref | ref |
| Quartiles2 | 1.86(1.26,2.75) | 0.002 | 1.74(1.15,2.65) | 0.009 | 3.48(1.53,7.91) | 0.003 | 3.27(1.37,7.76) | 0.007 |
| Quartiles3 | 2.01(1.37,2.95) | <0.001 | 1.91(1.26,2.88) | 0.002 | 3.62(1.61,8.12) | 0.002 | 3.38(1.45,7.88) | 0.005 |
| Quartiles4 | 4.11(2.85,5.92) | <0.001 | 3.09(2.04,4.67) | <0.001 | 8.91(4.16,19.1) | <0.001 | 6.41(2.79,14.7) | <0.001 |
| Per 1-mg/dl increased | 1.02(1.01,1.03) | <0.001 | 1.01(1.00,1.02) | <0.001 | 1.01(1.00,1.02) | <0.001 | 1.01(1.00,1.01) | 0.003 |
| **Combined GFR with microalbuminuria** |  |  |  |  |  |  |  |  |
| GFR>90, albuminuria(-) | ref | ref | ref | ref | ref | ref | ref | ref |
| GFR>90, albuminuria(+) | 1.67(1.14,2.44) | 0.008 | 1.55(1.02,2.35) | 0.038 | 2.89(1.56,5.34) | 0.001 | 2.87(1.47,5.62) | 0.002 |
| GFR>60, albuminuria(-) | 1.07(0.77,1.50) | 0.670 | 1.14(0.79,1.63) | 0.493 | 1.72(0.94,3.14) | 0.080 | 1.88(0.97,3.64) | 0.061 |
| GFR>60, albuminuria(+) | 2.51(1.78,3.53) | <0.001 | 2.40(1.64,3.53) | <0.001 | 4.06(2.25,7.33) | <0.001 | 3.82(1.96,7.47) | <0.001 |
| CKD, albuminuria(-) | 1.69(0.85,3.36) | 0.137 | 1.47(0.65,3.33) | 0.350 | 5.34(1.99,14.3) | 0.001 | 5.56(1.79,17.3) | 0.003 |
| CKD, albuminuria(+) | 3.07(1.87,5.05) | <0.001 | 3.37(1.91,5.95) | 0.001 | 6.40(2.98,13.8) | <0.001 | 6.41(2.64,15.6) | <0.001 |

Abbreviations: DR = diabetic retinopathy; VTDR= vision-threatened diabetic retinopathy; GFR = estimated glomerular filtration rate; OR=odds ratio; 95%CI= 95%confidence interval; SD=stand deviation
\*Adjusted for age and gender
†Adjusted for age, gender, diabetes duration, hypertension, HbA1c, MAP, BMI, use of insulin, CPR, TC and TG

To further assess the influence of microalbuminuria on the relationship between GFR and the presence of DR and VTDR, participants were divided into six groups according to different GFR levels and the presence of microalbuminuria. Our results showed that a low GFR was associated with any DR only in the presence of microalbuminuria (OR=2.40 for participants with GFR>60 and microalbuminuria (+), P<0.001; OR=3.37 for participants with CKD and microalbuminuria (+), P=0.001), while the GFR was an independent risk factor for VTDR regardless of whether microalbuminuria was present (all P<0.05) (Table 4).

Although a low GFR (OR=0.98 per 1 mL/min/1.73 m^2^ increase, P=0.007) and CKD (OR=2.95, P=0.011) were associated with DME in the age- and gender-adjusted logistic model, this association disappeared after adjusting for additional confounders. A significant association between DME and microalbuminuria was found in the third and fourth microalbuminuria quartiles in the multivariate logistic model, and this association increased from an OR of 2.99 in the third quartile (P=0.029) to an OR of 4.74 in the fourth quartile (P=0.002) (Table 5).

**Table 5.** Logistic regression of GFR and microalbuminuria with diabetic macular edema

| Factors | Model 1<sup>*</sup> |  | Model 2<sup>†</sup> |  |
| --- | --- | --- | --- | --- |
|  | OR(95%CI) | P value | OR(95%CI) | P value |
| **GFR(ml/min/1.73 m<sup>2</sup>)** |  |  |  |  |
| Normal, ≥90 | ref | ref | ref | ref |
| Mildly impaired, 60~89 | 1.33(0.75,2.35) | 0.332 | 1.18(0.62,2.25) | 0.605 |
| CKD, <60 | 2.95(1.29,6.75) | 0.011 | 2.33(0.88,6.16) | 0.088 |
| Per 1-ml/min/1.73m<sup>2</sup> increased | 0.98(0.97,0.99) | 0.007 | 0.99(0.97,1.01) | 0.085 |
| **Microalbuminuria(mg/dL)** |  |  |  |  |
| Quartiles1 | ref | ref | ref | ref |
| Quartiles2 | 1.81(0.64,5.16) | 0.266 | 1.82(0.62,5.32) | 0.276 |
| Quartiles3 | 3.22(1.25,8.30) | 0.015 | 2.99(1.12,7.95) | 0.029 |
| Quartiles4 | 5.89(2.39,14.6) | <0.001 | 4.74(1.79,12.6) | 0.002 |
| Per 1-mg/dl increased | 1.00(0.99,1.01) | 0.060 | 1.01(0.99,1.01) | 0.348 |

Abbreviations: GFR = estimated glomerular filtration rate; OR=odds ratio;
95%CI= 95%confidence interval; SD=stand deviation
<sup>\*</sup>Adjusted for age and gender
<sup>†</sup>Adjusted for age, gender, diabetes duration, hypertension, HbA1c, MAP, BMI, use of insulin, CPR, TC and TG

## Discussion

In the current study, we found that both a low GFR level and a high level of microalbuminuria were significantly associated with the presence of any DR and VTDR in Chinese T2DM participants. We further assessed the influence of microalbuminuria on the association of the GFR with the presence of DR, and the results showed that a low GFR was associated with any DR only in the presence of microalbuminuria, while the GFR was an independent risk factor for VTDR even without presence of microalbuminuria. The microalbuminuria was an independent risk factor for the presence of DME.

The exact relationship between the GFR and DR is still controversial, with some studies demonstrating a strong negative association between the GFR and DR^10, 11, 24-27^ and other studies indicating no association or an indirect association.^12, 20, 28^ Some studies indicated an association between GFR and DR only under certain scenarios. The results of the National Health and Nutrition Examination Survey (NHANES) showed a significant association between DR and a decreased GFR in non-Hispanic Blacks, those with a BMI≥30 kg/m^2^, or those who were not taking a renin-angiotensin-aldosterone system (RAAS) inhibitor.^26^ Other studies indicated that there were racial differences in the associations between GFRs and DR.^11^ Our study indicated that a low GFR level was significantly associated with an increased presence of DR and VTDR. Additionally, we found that a low GFR was associated with any DR only in the presence of microalbuminuria, while the GFR was independently associated with VTDR even without the presence of microalbuminuria. One study from Italy consistently showed an association between a low GFR and advanced DR regardless of whether microalbuminuria was present, although the association was strongest in the microalbuminuria group.^25^ While the Singapore Prospective Study Program showed that low GFR levels were associated with any DR and advanced DR only in the presence of microalbuminuria.^20^ Our study and previous studies further confirmed the hypothesis of shared pathogenesis mechanisms involving microvascular damage to retinal and renal vessels, leading to progression to DR and DN in diabetic patients.^29^ Further longitudinal studies should be performed to verify the exact causal relationship between GFR and DR in future.

Many studies have investigated the association between microalbuminuria and DR, with some studies indicating a strong association^12, 30, 31^ and others not.^26^Studies evaluating this association in Chinese diabetic patients are limited. One study from Shanghai indicated that microalbuminuria was an independent risk factor for DR.^28^ The present study consistently showed that a high microalbuminuria level was a strong risk factor for any DR and VTDR. In addition to being a risk factor for renal damage, microalbuminuria was also associated with other diabetic micro- and macrovascular complications, suggesting its role as an indicator of generalized vascular damage.^32^ Thus, the close monitoring of microalbuminuria may help clinicians detect and treat DR early.

Existing studies on the relationship between microalbuminuria and DME are limited. One study from Korea showed that microalbuminuria was not associated with DME in 971 Korean DM patients.^27^ Other studies have shown a positive relationship between microalbuminuria and DME.^33^ Our study also indicated that microalbuminuria was significantly associated with DME, and the risk increased as microalbuminuria increased. However, we should interpret these results with caution because this association was significant only in the categorical variable analysis but not in the continuous variable analysis. The small number of participants with DME may partly explain our results that the association between microalbuminuria and DME may have occurred by chance. Further studies on DME and microalbuminuria should be conducted with different races and disease stage severities.

The reported disparity of association between DR and DN may be partly due to differences in study designs, subject races and study sample sizes.

Additionally, the current diagnosis of DN is based on clinical manifestations other than renal biopsy. Thus, it is impossible to distinguish DN from nondiabetic renal disease, for which the incidence rate varies from 27% to 82.9% among DM patients. Current studies on the pathophysiology of renal lesions and DR are limited.^34, 35^ One study of 250 diabetic participants with biopsy-proven DN at West China Hospital found that the severity of glomerular lesions was significantly associated with DR and that DR was an independent risk factor for renal change progression.^34^ Further studies are needed to investigate the association of DR with DN and the exact mechanism.

The strengths of this study include the large sample size, detailed ocular examinations with standardized protocol. Each participant underwent 7-field fundus imaging, which ensured the possible maximal grade for the presence of DR, as some features of DR occurred in the peripheral retina, which may be missed in 2-field fundus imaging. The main limitation is that our study involved community-based recruitment from the Yuexiu Family Doctor Project, and selection bias may exist. Thus, the study conclusions may not be generalizable to the general population. However, the relatively large study population obtained by consecutive recruitment made the current study reliable. Second, we used the UK NDESP grading system instead of the commonly used ETDRS grading system which comprises different definitions. However, these two grading systems performed similarly when used to assess the presence and severity of DR, thus having little effect on the results.

In conclusion, the current study indicated that a low GFR was associated with the presence of any DR only in the presence of microalbuminuria, while the GFR was independently associated with VTDR regardless of microalbuminuria status among Chinese diabetic participants. A high level of microalbuminuria was significantly associated with the presence and severity of DR as well as the presence of DME. Our findings may provide evidence of the potential mechanisms underlying DR and DN and have clinical implications for the management of DM. Further longitudinal studies on the association between renal function and DR are needed to confirm the causal relationship and potential mechanisms.

## Data Availability

The availability of all data referred to in the manuscript and note links below.

## References

1. Wild S, Roglic G, Green A, Sicree R, King H. Global prevalence of diabetes: estimates for the year 2000 and projections for 2030. Diabetes care 2004; 27(5): 1047–53.

2. Yau JW, Rogers SL, Kawasaki R, et al. Global prevalence and major risk factors of diabetic retinopathy. Diabetes care 2012; 35(3): 556–64.

3. Echouffo-Tcheugui JB, Ali MK, Roglic G, Hayward RA, Narayan KM. Screening intervals for diabetic retinopathy and incidence of visual loss: a systematic review. Diabetic medicine : a journal of the British Diabetic Association 2013; 30(11): 1272–92.

4. Younis N, Broadbent DM, Vora JP, Harding SP, Liverpool Diabetic Eye S. Incidence of sight-threatening retinopathy in patients with type 2 diabetes in the Liverpool Diabetic Eye Study: a cohort study. Lancet 2003; 361(9353): 195–200.

5. Flaxman SR, Bourne RRA, Resnikoff S, et al. Global causes of blindness and distance vision impairment 1990-2020: a systematic review and meta-analysis. The Lancet Global health 2017; 5(12): e1221–e34.

6. Zheng Y, He M, Congdon N. The worldwide epidemic of diabetic retinopathy. Indian journal of ophthalmology 2012; 60(5): 428–31.

7. Gonzalez Blanco F, Sanz Fernandez JC, Munoz Sanz MA. Axial length, corneal radius, and age of myopia onset. Optometry and vision science : official publication of the American Academy of Optometry 2008; 85(2): 89–96.

8. Barrett EJ, Liu Z, Khamaisi M, et al. Diabetic Microvascular Disease: An Endocrine Society Scientific Statement. The Journal of clinical endocrinology and metabolism 2017; 102(12): 4343–410.

9. Dronavalli S, Duka I, Bakris GL. The pathogenesis of diabetic nephropathy. Nature clinical practice Endocrinology & metabolism 2008; 4(8): 444–52.

10. Kaewput W, Thongprayoon C, Rangsin R, Ruangkanchanasetr P, Mao MA, Cheungpasitporn W. Associations of renal function with diabetic retinopathy and visual impairment in type 2 diabetes: A multicenter nationwide cross-sectional study. World journal of nephrology 2019; 8(2): 33–43.

11. Man RE, Sasongko MB, Wang JJ, et al. The Association of Estimated Glomerular Filtration Rate With Diabetic Retinopathy and Macular Edema. Investigative ophthalmology & visual science 2015; 56(8): 4810–6.

12. Chen YH, Chen HS, Tarng DC. More impact of microalbuminuria on retinopathy than moderately reduced GFR among type 2 diabetic patients. Diabetes care 2012; 35(4): 803–8.

13. Perkins BA, Ficociello LH, Silva KH, Finkelstein DM, Warram JH, Krolewski AS. Regression of microalbuminuria in type 1 diabetes. The New England journal of medicine 2003; 348(23): 2285-93.

14. Hovind P, Tarnow L, Rossing P, et al. Predictors for the development of microalbuminuria and macroalbuminuria in patients with type 1 diabetes: inception cohort study. Bmj 2004; 328(7448): 1105.

15. Macisaac RJ, Ekinci EI, Jerums G. Markers of and risk factors for the development and progression of diabetic kidney disease. American journal of kidney diseases : the official journal of the National Kidney Foundation 2014; 63(2 Suppl 2): S39–62.

16. Kramer HJ, Nguyen QD, Curhan G, Hsu CY. Renal insufficiency in the absence of albuminuria and retinopathy among adults with type 2 diabetes mellitus. Jama 2003; 289(24): 3273–7.

17. Retnakaran R, Cull CA, Thorne KI, Adler AI, Holman RR, Group US. Risk factors for renal dysfunction in type 2 diabetes: U.K. Prospective Diabetes Study 74. Diabetes 2006; 55(6): 1832–9.

18. Mottl AK, Kwon KS, Mauer M, Mayer-Davis EJ, Hogan SL, Kshirsagar AV. Normoalbuminuric diabetic kidney disease in the U.S. population. Journal of diabetes and its complications 2013; 27(2): 123–7.

19. MacIsaac RJ, Tsalamandris C, Panagiotopoulos S, Smith TJ, McNeil KJ, Jerums G. Nonalbuminuric 15. renal insufficiency in type 2 diabetes. Diabetes care 2004; 27(1): 195–200.

20. Sabanayagam C, Foo VH, Ikram MK, et al. Is chronic kidney disease associated with diabetic retinopathy in Asian adults? Journal of diabetes 2014; 6(6): 556–63.

21. Revised Grading Definitions for the NHS Diabetic Eye Screening Programme. Available at: https://www.gov.uk/government/publications/diabetic-eye-screening-retinal-imagegrading-criteria. Accessed June 19, 2015.

22. Levey AS, Stevens LA, Schmid CH, et al. A new equation to estimate glomerular filtration rate.Annals of internal medicine 2009; 150(9): 604–12.

23. Kdoqi. KDOQI Clinical Practice Guidelines and Clinical Practice Recommendations for Diabetes and Chronic Kidney Disease. American journal of kidney diseases : the official journal of the National Kidney Foundation 2007; 49(2 Suppl 2): S12–154.

24. Rodriguez-Poncelas A, Mundet-Tuduri X, Miravet-Jimenez S, et al. Chronic Kidney Disease and Diabetic Retinopathy in Patients with Type 2 Diabetes. PloS one 2016; 11(2): e0149448.

25. Penno G, Solini A, Zoppini G, et al. Rate and determinants of association between advanced retinopathy and chronic kidney disease in patients with type 2 diabetes: the Renal Insufficiency And Cardiovascular Events (RIACE) Italian multicenter study. Diabetes care 2012; 35(11): 2317–23.

26. Mottl AK, Kwon KS, Garg S, Mayer-Davis EJ, Klein R, Kshirsagar AV. The association of retinopathy and low GFR in type 2 diabetes. Diabetes research and clinical practice 2012; 98(3): 487–93.

27. Lee WJ, Sobrin L, Lee MJ, Kang MH, Seong M, Cho H. The relationship between diabetic retinopathy and diabetic nephropathy in a population-based study in Korea (KNHANES V-2, 3). Investigative ophthalmology & visual science 2014; 55(10): 6547–53.

28. Chen H, Zheng Z, Huang Y, et al. A microalbuminuria threshold to predict the risk for the development of diabetic retinopathy in type 2 diabetes mellitus patients. PloS one 2012; 7(5): e36718.

29. Romero-Aroca P, Mendez-Marin I, Baget-Bernaldiz M, Fernendez-Ballart J, Santos-Blanco E. Review of the relationship between renal and retinal microangiopathy in diabetes mellitus patients. Current diabetes reviews 2010; 6(2): 88–101.

30. Girach A, Vignati L. Diabetic microvascular complications--can the presence of one predict the development of another? Journal of diabetes and its complications 2006; 20(4): 228–37.

31. Ajoy Mohan VK, Nithyanandam S, Idiculla J. Microalbuminuria and low hemoglobin as risk factors for the occurrence and increasing severity of diabetic retinopathy. Indian journal of ophthalmology 2011; 59(3): 207–10.

32. Savage S, Estacio RO, Jeffers B, Schrier RW. Urinary albumin excretion as a predictor of diabetic retinopathy, neuropathy, and cardiovascular disease in NIDDM. Diabetes care 1996; 19(11): 1243–8.

33. El-Asrar AM, Al-Rubeaan KA, Al-Amro SA, Moharram OA, Kangave D. Retinopathy as a predictor of other diabetic complications. International ophthalmology 2001; 24(1): 1–11.

34. Zhang J, Wang Y, Li L, et al. Diabetic retinopathy may predict the renal outcomes of patients with diabetic nephropathy. Renal failure 2018; 40(1): 243–51.

35. Olsen S, Mogensen CE. How often is NIDDM complicated with non-diabetic renal disease? An analysis of renal biopsies and the literature. Diabetologia 1996; 39(12): 1638–45.

